# PREPRINT: Mechanisms of Motoric Cognitive Risk – hypotheses based on a systematic review and meta-analysis of longitudinal cohort studies of older adults

**DOI:** 10.1101/2021.10.26.21265519

**Authors:** Donncha S. Mullin, Alastair Cockburn, Miles Welstead, Michelle Luciano, Tom C. Russ, Graciela Muniz-Terrera

## Abstract

We aimed to refine the hypothesis that Motoric Cognitive Risk (MCR), a syndrome combining measured slow gait speed and self-reported cognitive complaints, is prognostic of incident dementia and other major causes of morbidity in older age. We propose mechanisms on the relationship between motor and cognitive function and describe a roadmap to validate these hypotheses. We systematically searched major electronic databases from inception to August 2021 for original longitudinal cohort studies of adults aged ≥60 years that compared an MCR group to a non-MCR group with any health outcome. Fifteen cohorts were combined by meta-analysis. Participants with MCR were at an increased risk of cognitive impairment (adjusted hazard ratio [aHR] 1.76, 95%CI 1.49-2.08; I^2^=24.9%), dementia (aHR 2.12, 1.85-2.42; 33.1%), falls (adjusted Relative Risk 1.38, 1.15-1.66; 62.1%), and mortality (aHR 1.49, 1.16-1.91; 79.2%). The prognostic value of MCR is considerable and mechanisms underlying the syndrome are proposed.

**Systematic review registration:** PROSPERO CRD42020225183.

## 1. Narrative

### 1.1 Central question

Can a gait-based syndrome, Motoric Cognitive Risk (MCR), predict dementia and other age-related negative health outcomes? If so, what are the possible underlying mechanisms?

### 1.2 Objective

This paper is a proposal for an update of the Hypothesis on the Motoric-cognitive Mechanism in Neurodegeneration-Dementia-Alzheimer^1^ syndrome (abbreviated hereafter as the Hypothesis) based on a thorough systematic review and meta-analysis of the evidence. The present work intends to: (1) promote new thinking about the prognostic value of MCR in dementia and other major causes of morbidity in older age; and (2) propose shared mechanisms between MCR and neurodegeneration and outline a roadmap for further work to validate these hypotheses. This report aims to synthesize and critique current evidence on MCR and describe challenges to the MCR concept and the use of MCR as a clinical tool.

### 1.3 Current knowledge

As there are still no effective treatments for dementia, any biomarker that supports early identification of high-risk individuals would allow time for lifestyle modification, planning for future care needs, and could ultimately contribute to a reduction in overall prevalence of dementia. Slow gait speed and self-reported cognitive complaints show potential to be such biomarkers as they are some of the earliest reported precursors in the pre-clinical stage of dementia, occurring 10-15 years before dementia diagnosis.^1,2^ Motoric cognitive risk (MCR) syndrome is a high-risk state combining objective (measured) slow walking speed and subjective (self-reported) cognitive complaint in those able to ambulate and in the absence of dementia.^3^

First defined by Verghese in 2013^4^, MCR has incremental predictive ability over either slow gait or cognitive complaint alone.^5^ Gait is a complex task requiring coordination between widespread brain regions, therefore MCR may reflect neurodegeneration occurring in the preclinical stage of dementia. Imaging studies in the MCR field indicate atrophy of executive function areas (frontal and pre-frontal lobe networks).^6^

The only existing meta-analysis of MCR^7^ found that MCR predicts cognitive impairment and dementia. However, this meta-analysis had significant methodological limitations, including pooling the results of different effect measures (e.g., hazard ratios and odds ratios) and, focusing on the term “MCR”, thus only including studies published from 2013 onward and excluding earlier papers examining the same construct without naming it as such. We address these limitations in our study. Other studies report that MCR identifies those at risk of falls,^8–10^ post-falls hip fractures,^9^ disability,^11^ and mortality,^12,13^ but these studies have not yet been pooled in a meta-analysis. Other non-systematic MCR reviews focused on risk factors for developing MCR^14^ or aimed to give a more general overview of the MCR construct.^6^

### 1.4 Knowledge gap and importance of this study

Important conditions such as dementia or falls are projected to affect such a large number of people over the next 30 years that even small reductions in the incidence – or delaying the age of onset which would have the same result – are likely to have significant effects on numbers of people affected and consequently the huge associated public health costs.^5,14^ Could there be shared mechanisms explaining the association of MCR with both these outcomes? If so, identifying and targeting these mechanisms could reduce the prevalence of these major causes of morbidity and mortality in older adults. Furthermore, if a quick, inexpensive, and easy-to-measure clinical construct could reliably identify people at high risk of developing either or both dementia and falls, along with other adverse health outcomes such as cognitive impairment and mortality, it would be an important public health tool and would be equally implementable in low-to-middle income countries. Our study addresses this knowledge gap and proposes hypotheses on the underlying mechanisms.

### 1.5 Limitations

We were unable to obtain raw data from every eligible study to allow for calculation of a comparable summary effect measure for each study for inclusion in the meta-analysis – an individual participant meta-analysis. Therefore, some studies had to be left out of the pooled result, reducing the overall power. We believe this important compromise ensures our findings are as valid and reliable as possible based on the published literature while avoiding potential significant delays to undertaking this novel work. Most studies in our review had a degree of risk of bias (ROB) due to how they managed missing data and confounding factors, and a lack of generalizability (section 2.1.6). These are common limitations in cohort studies and partly explain why they are lower down the hierarchy of evidence compared to randomized controlled trials. We accounted for substantial heterogeneity in the mortality outcome by downgrading our Grading of Recommendations Assessment, Development and Evaluation (GRADE) certainty assessment for inconsistency (see section 2.2.5). Although some might avoid pooling our mortality data due to statistical heterogeneity, we believe the summary data provide an important global perspective on MCR.^13^ There is a relatively small number of studies for each health outcome in the meta-analysis, meaning that statistical tests for publication bias lacked power to detect real asymmetry from chance, although visual inspection of funnel plots is reassuring (section 2.3.5). Finally, dementia is a clinical diagnosis, but in the studies included it was often determined using surrogate markers such as scores in cognitive tests, increasing heterogeneity and decreasing the generalizability of our findings. This is a common issue in dementia research using cohort data. Reassuringly, the results of studies that diagnosed dementia using clinical criteria^4,5^ were consistent with those using surrogate markers.^11,15^

1.6 Discussion

Our results for risk of cognitive impairment and dementia in participants with MCR at baseline were consistent with the only existing meta-analysis on the topic.^7^ This earlier meta-analysis reported that individuals with MCR were at a 70% increased risk of developing cognitive impairment (adjusted Hazard Ratio [aHR] 1.70, 95%CI 1.46-1.98) and a 150% increased risk of developing dementia (aHR 2.50, 95%CI 1.75-2.39). The slight differences reflect our inclusion of new studies and two major strengths of our study, namely our decisions to only pool summary effect measures of the same type and to always use the most adjusted effect measure reported. Accordingly, our findings have reduced confounding and are likely to be more conservative. Furthermore, by focusing on the term “MCR”, the previous meta-analysis only included studies published from 2013 onward, unlike our search which captured any study combining slow gait and subjective cognitive complaint since database inception.

Our paper makes methodological advances on the existing meta-analysis with regards to cognitive impairment and dementia outcomes,^7^ and it is the first to meta-analyze MCR studies reporting on falls and mortality outcomes. As MCR is a recently defined construct, more research is needed to increase certainty in each of our findings, particularly falls prediction. Other outcomes more likely in those with MCR at baseline include recurrent falls, post-falls hip fractures, and disability, but these outcomes require further research to allow for robust meta-analysis.

#### 1.6.1 Hypotheses on mechanisms underlying MCR

Interactions between MCR, poor brain health, falls, and increased mortality are likely due to a range of biological, psychological, and social mechanisms. Causality is likely to be bidirectional, and the mechanisms may be multifactorial. There is unlikely to be one, unifying mechanism linking MCR with these negative health outcomes, but, based on recent evidence, we propose hypotheses to explain some of the potential underlying biological mechanisms.

##### Epidemiological and clinical

MCR prevalence increases with age with rates of 8.9% in the 60-74 years group and 10.6% in the ≥75years group^5^. MCR may increase the risk of dementia, falls and mortality by contributing to geriatric syndromes such as delirium, depression, and medication mismanagement. Lower education is associated with increased risk of MCR^5,13,16^ and is an established risk factor for dementia. Low physical activity is another lifestyle risk factor shared by MCR with dementia, falls, disability, and increased mortality.^11,13,17^ Reduced concentration and psychomotor retardation are well-recognized symptoms of depression, so it is no surprise that depression has been associated with MCR.^18^ Personality traits such as neuroticism^19^ have also been associated with MCR, which may in part be due to an increased likelihood in this population to report subjective cognitive complaint. Pilot trials to improve executive function by cognitive training, dual-task ‘walking while talking’, or brain stimulation have improved gait speed.^6^

##### Neuropathology

Emerging data suggest that both neurodegenerative and vascular changes may contribute to progression to dementia in those with MCR.^20^ It is widely accepted that cerebral small vessel disease (SVD) is an important cause of dementia and it is estimated that over one third of dementias could be prevented by preventing stroke.^21^ Lacunar infarcts in the frontal lobe were associated with MCR even after adjusting for vascular risk factors and presence of white matter hyperintensities (adjusted Odds Ratio (aOR): 4.67, 95% CI: 1.69-12.94).^22^ SVD is also a significant contributor to risk of falling and SVD, especially in the frontal lobes, and has been linked to increased mortality.^12^ However, adults free of dementia with slow gait had associated amyloid β brain deposition, independent of underlying vascular change.^23^

Cognition and gait share many other risk factors such as cardiovascular disease and diabetes mellitus^24^ so it is no surprise that a pooled meta-analysis found that MCR was associated with both of these chronic conditions, as well as hypertension and stroke.^25^ These findings support our hypothesis that a vascular mechanism may underlie the pathophysiology of MCR syndrome.

##### Neuroimaging and neurophysiology

A 2019 review reported that MCR was associated with lower gray matter volume in the premotor and prefrontal cortices, but had no significant association with white matter abnormalities.^7^ The authors concluded that the pathophysiology of MCR was more likely due to neurodegenerative rather than ischemic lesions.^7^ This conclusion was admittedly based on a small number of imaging studies and it was hypothesized that MCR detects individuals at such an early stage of the disease process leading to dementia that the consequences of the vascular component may not yet be detected.^7^ Furthermore, MCR was associated with frontal lacunar infarcts in a study of 139 older adults in India.^22^

##### Genetics

The first study to investigate individual-level genetic burden in relation to a predementia syndrome examined the polygenic inheritance of MCR in a sample of 4,915 older individuals.^26^ The authors examined nine phenotypes associated with MCR and found that obesity-related genetic traits increase the risk of MCR syndrome. Obesity in older adults is an established risk factor in falls, disability and increased mortality.^27,28^

A prospective examination of inflammatory cytokine genes found that polymorphisms which lead to over-expression of the anti-inflammatory cytokine Interleukin-10 (IL-10) are associated with increased MCR incidence.^29^ While this makes any shared neuroinflammatory pathway between MCR and dementia less probable, an over-expression of IL-10 points toward a proamyloidogenic hypothesis of cognitive decline.^29^ Early work in mice models found that IL-10 expression leads to increased amyloid β accumulation and reduction in synaptic proteins as well as increasing expression of *APOE* and suppressed phagocytosis of β-amyloid by microglia.^30^ This link between *APOE* and MCR is perhaps unsurprising as the *APOE* ε4 allele was independently associated with increased risk of gait speed decline and disability in older men.^31^

##### Cellular mechanisms

No specific mechanistic work on MCR at the cellular level has been performed to date. However, emerging evidence on the effects of Alzheimer’s disease pathology on motor neuron function in transgenic mice merit consideration. For example, many studies have reported on the impaired motor performance such as beam walking and significant motor neuron axonopathy in transgenic mice with β-amyloid mutations.^1,32^ It will be fascinating to assess if future studies treating developed or preventing Alzheimer’s disease pathologies improves or prevents further declines of motor function.^1^

### 1.7 Major challenges for the Hypothesis

First, the definition of MCR requires standardization to allow for better comparison of MCR prevalence rates and prognostic value across populations. The range of methods for diagnosing subjective cognitive complaint highlights the need for a consistent definition of MCR in future studies.

Early work by Verghese et al.^4^ found that MCR was better at predicting vascular dementia than Alzheimer dementia, but more recent work in an independent cohort^15^ found conversely that MCR was better at predicting Alzheimer’s dementia than non-Alzheimer’s dementia. This challenges the premise that early upstream vascular alterations degrade the brain structures shared by cognitive and motor function systems, namely frontal and prefrontal motor cortex.^20,21^

### 1.8 Conclusions and next steps

The motoric-cognitive hypothesis of neurodegeneration does not seem to have one unifying underlying mechanism, so future steps to elucidate these will require a multidisciplinary approach exploring the issue using complementary techniques.

1. The relative contributions from cortical motor regions to neuropsychological tests that comprise part of the clinical diagnosis of dementia remain to be addressed.^1^
2. Longitudinal studies of at-risk populations with genetic, neurophysiological, neuroimaging, or other biomarkers and pathological validation will help determine the progress of motor and cognitive impairment in dementia.^33^
3. A large-scale discovery genome-wide association study of MCR is an important step to identify underlying biological mechanisms of MCR. This would identify targets for further investigation and possibly treatment to reduce MCR and ultimately dementia, falls, disability, and excess mortality. The generation of a reliable MCR polygenic risk score might have clinical utility for early prediction, and thus prevention, of those at risk of MCR.
4. Interventional trials are important to assess whether targeting slow gait and subjective cognitive impairment delays or reverses MCR, and whether this reduces transition to cognitive impairment, dementia, falls, and excess mortality.

## 2. Consolidated Results and Study Design

### 2.1 Methodology

This systematic review was conducted following the updated guidelines of Preferred Reporting Items for Systematic reviews and Meta-Analyses (PRISMA 2020)^34^ and the meta-analysis followed the Meta-Analyses of Observational Studies in Epidemiology (MOOSE) guidelines.^35^ The protocol was pre-registered in the international prospective register of systematic reviews (PROSPERO CRD42020225183).

#### 2.1.1 Search strategy

We searched the AMED, APA PsychInfo, CINAHL, Cochrane, EMBASE and Ovid MEDLINE databases from conception to 20/08/2021, then carried out a backwards and forwards citation search with no language or publication date restrictions. The search strategy was devised iteratively with support from an academic librarian, and the strategy was peer reviewed using the Peer Review of Electronic Search Strategies (PRESS) checklist.^36^ The full search strategy is in supplementary table 1.

##### 2.1.2 Eligibility criteria

We used the PICOT (Population; Intervention; Comparator; Outcome; Timing/Type) system to design our review question (supplementary box 1).^37^ In summary, we examined longitudinal cohort studies of community-based adults aged ≥60 years with an MCR group compared to a non-MCR group for any health outcome with a minimum of one year follow-up. Only peer-reviewed full-text articles were included in the meta-analysis and synthesis.

##### 2.1.3 Screening and selection

Two authors (DM and AC) independently reviewed all titles then all included abstracts using Covidence software.^38^ If the study appeared to meet the selection criteria, the same two investigators independently reviewed the full text. Discrepancies were resolved through open discussion and verified by a third author when necessary.

##### 2.1.4 Data collection process

We iteratively designed a bespoke data extraction tool based on the CHARMS-PF (CHecklist for critical Appraisal and data extraction for systematic Reviews of prediction Modelling Studies, adapted for prognostic factors) template^37^, which two authors (DM and AC) used independently to extract data from eligible studies (supplementary table 2). We then compared the extracted data and resolved any discrepancies through discussion and referring to the study in question. The combined data extraction tool was then double-checked for accuracy. Whenever study details were unclear, we contacted study authors for further information.

###### 2.1.5 Data items

We extracted the following data for the exposure variable (MCR): slow gait measurement protocol, average gait speed, subjective cognitive complaint measurement method and MCR prevalence rate. For our outcome variables, we recorded method of measurement as well as the most adjusted model results of any health outcome result, whether reported as adjusted hazard ratio (aHR), adjusted odds ratio (aOR), or adjusted relative risk (aRR), and their corresponding 95% confidence intervals (CI) and p-values, if available. Unadjusted model results were not reported frequently enough to allow meaningful comparison on synthesis. To inform assessment of residual confounding, we recorded the covariates adjusted for in each model. Where an outcome was reported over various follow-up timepoints we selected those timepoints most common across studies to minimize meta-analysis heterogeneity. Other data items extracted included, author name, year, country, cohort, size, the study design, and participant characteristics.

###### 2.1.6 Study quality and risk of bias assessment

We performed a ROB assessment, using an expanded version of the Quality In Prognosis Studies (QUIPS) tool recommended by the Cochrane Prognosis Methods Group to assess ROB in prognostic factor studies.^37^ We generated a summary ROB plot illustrating the overall ROB of the literature base, and a ROB traffic light plot to illustrate the ROB of each study. The ROB assessments were incorporated into our meta-analysis at the grading of evidence stage.

##### 2.2 Meta-analysis

###### 2.2.1 Eligibility for each synthesis

We tabulated the study outcome characteristics and compared each against our planned outcome to ensure that the study outcome was valid. We only included in the meta-analysis those studies with outcomes which were judged satisfactory by our clinical content experts. Only health outcomes reported from at least three cohorts were included in the meta-analysis, to ensure appropriate synthesis.

###### 2.2.2 Effect measures

We used aHR and 95% CI to synthesize studies reporting cognitive impairment, dementia, and mortality outcomes, and aRR and 95% CIs to synthesize studies reporting falls as the outcome. These were the effect measures most reported in eligible studies for each of these outcomes, thus allowing inclusion of most studies. When a study reported the effect measure in a way incompatible with our analysis, we contacted the authors to request data to allow for our own calculation (e.g., aHR) or we converted the effect measure (e.g., aOR to aRR), if possible and appropriate, based on methods suggested by Tierney.^39^ If these attempts were not successful, we omitted the study from our meta-analysis to avoid comparing different effect measures in the one analysis. In all cases, only the most adjusted effect measures were used.

###### 2.2.3 Synthesis methods

We log-transformed our effect measures and their 95% CIs to make them normally distributed, an assumption of our meta-analysis model. We then calculated log standard error using the methods described in the Cochrane Handbook v6.2.^40^

We used a random-effect model (REM) to allow for within-study sampling error and between-studies variability due to varying study characteristics. The relative lack of small studies supported our decision to use the REM approach (as smaller studies receive larger weights in REM in comparison to fixed-effect modelling). The degree of heterogeneity was calculated using a restricted maximum-likelihood estimator, following recent guidance.^40^ The extent and impact of between-study heterogeneity were visually displayed in forest plots and reported as the tau-squared and I-squared statistic, accompanied by 95% CIs to judge our confidence about these metrics. I-squared was chosen over Cochrane’s Q to better account for the small number of studies in each analysis. Prediction intervals are included in the forest plots to illustrate the range for which we can expect the effects of future studies to fall, based on our present evidence in the meta-analysis. Prediction intervals help overcome any limitations of the I-squared and tau-squared methods.^41^ Meta-analysis was performed by one author in R (version 4.0.2) using the metafor (2.4.0) and dmetar (0.0.9) packages.^42^

We tabulated our study characteristics structured by outcome domain, ordered from low to high ROB to orientate readers to the most robust evidence (table 1). The results of the meta-analyses are displayed in forest plots for each outcome, displaying the effect estimates and confidence intervals of each study and the summary estimate. The plots are ordered by study weight to highlight any patterns in the data.

**Table 1.**
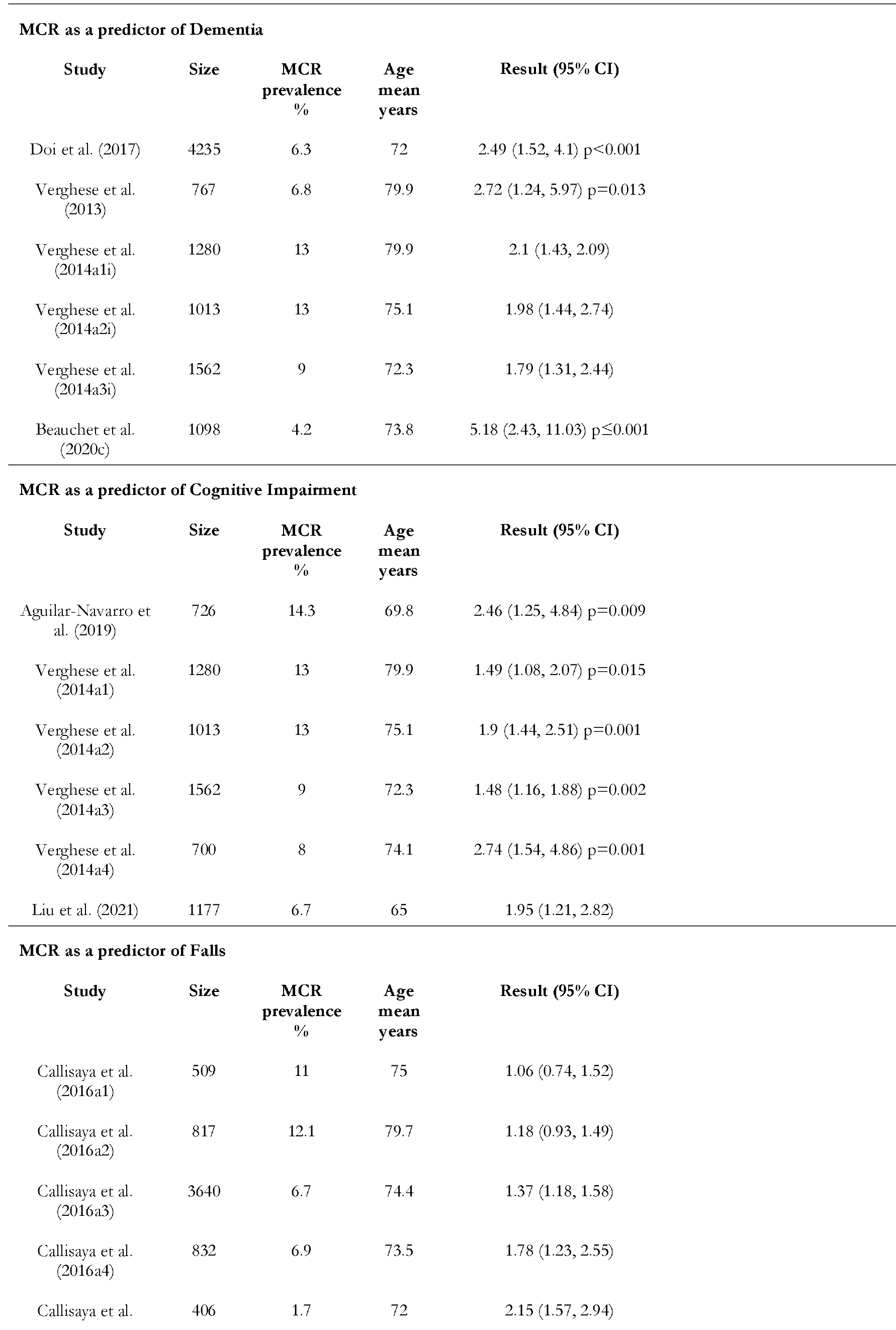
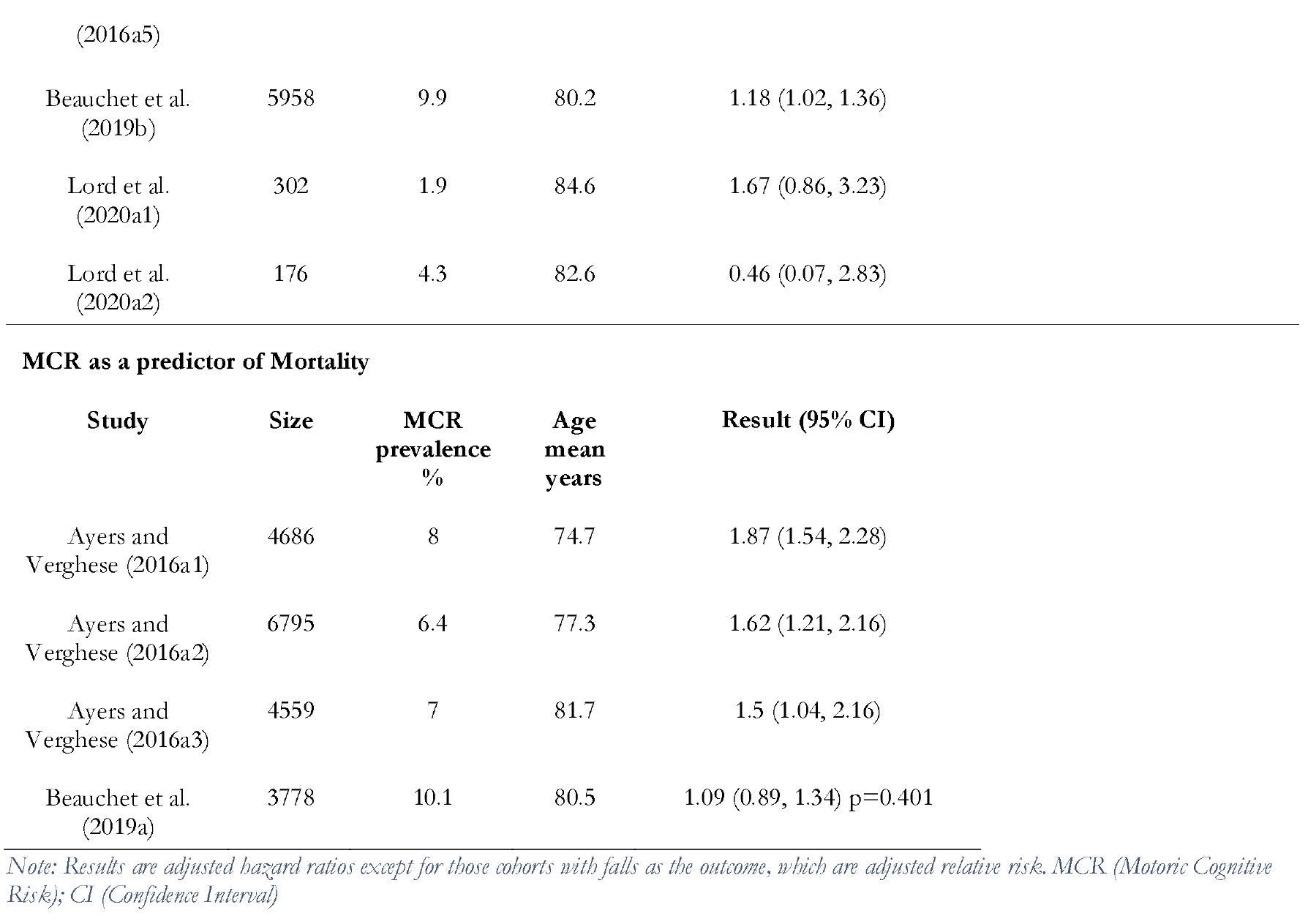
Characteristics and results of the studies included in the meta-analysis (further details in supplementary table 3)

##### 2.2.4 Assessing publication bias or outcome reporting bias

We produced funnel plots for meta-analysis of MCR for each health outcome to allow for visual assessment of small-study effects. These were assessed for asymmetry using Egger’s test. To assess outcome reporting bias, two reviewers independently compared the outcomes reported in the methods and results sections of the studies.

##### 2.2.5 Certainty assessment methods

Our default starting position regarding certainty in the level of evidence was in keeping with the Cochrane guidance to generally regard evidence from sound observational studies as low quality.^40^ We modified the GRADE considerations adapted for prognosis research as recommended by Huguet^43^ to assess the certainty of the body of evidence as it related to the studies included in the meta-analyses for each outcome. See section 3.1 and supplementary box 2 for full details.

#### 2.3 Results

##### 2.3.1 Study selection

We found 705 records on database searching, following de-duplication. We sought retrieval of full-text reports for 94. We included 15 studies from the databases search,^4,5,8–13,15–17,44–47^ with 11 of these proving eligible for meta-analysis. We then performed backwards and forwards citation searching by reviewing the reference list of these studies, as well as those of three reviews^7,14,48^ and one editorial paper^33^ on MCR. Of the 1800 records found this way, we reviewed full-text reports for 35 but included no additional papers. One longitudinal study which appeared to meet inclusion criteria was excluded as the MCR group defined using slow gait speed as a criteria was not followed up longitudinally.^3^ Figure 1 shows the PRISMA flow diagram of study selection.

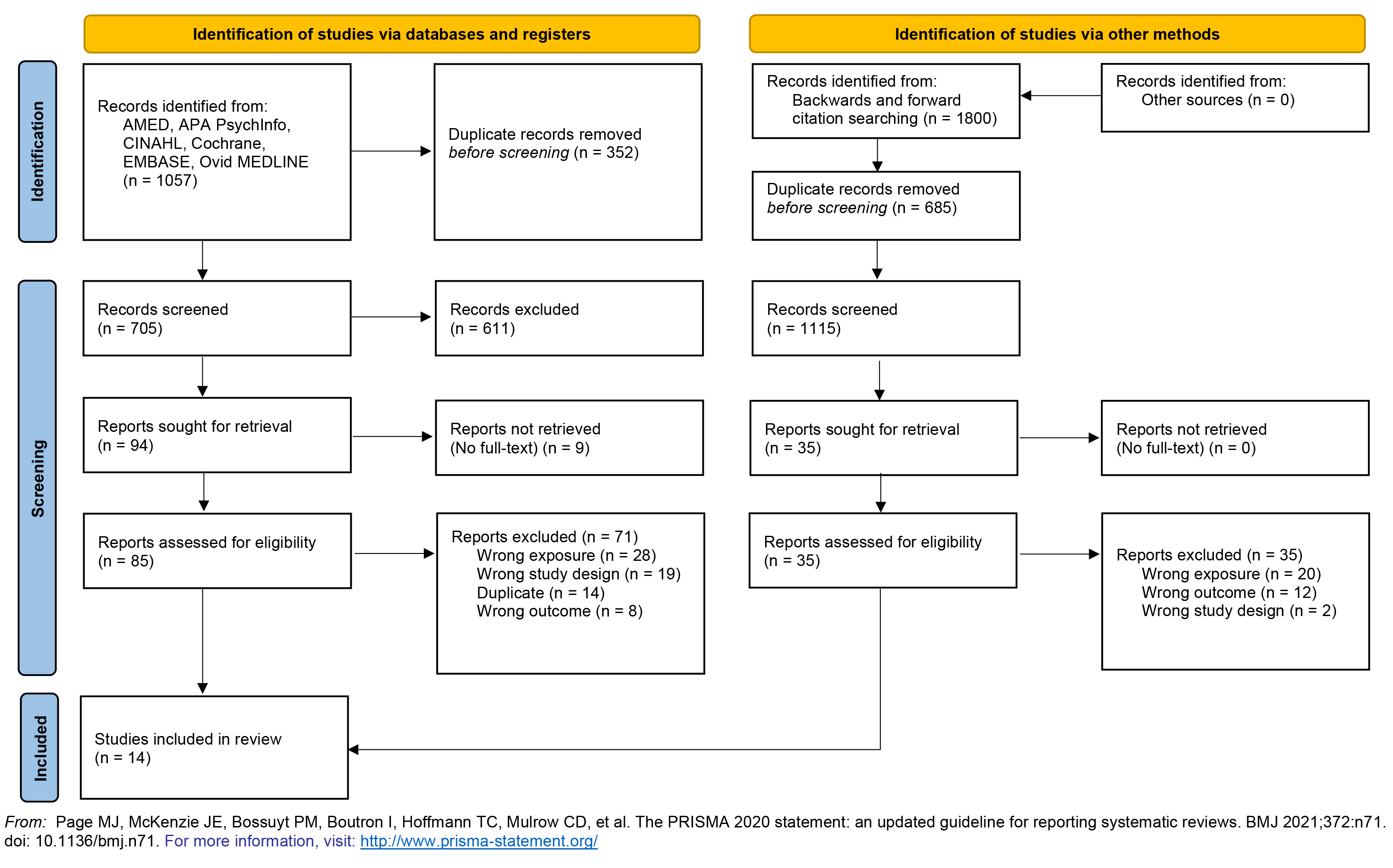

##### 2.3.2 Study characteristics

###### Error! Reference source not found

shows the key characteristics for each cohort included in the meta-analysis, structured by outcome, and ordered by ROB from low to high risk. More detail is available in supplementary table 3. All studies were published from 2013 onwards. Six cohorts were based in the USA, three from European countries, and one from each of Canada, Japan, China, Mexico, Australia, and New Zealand. Cohort sizes ranged from 176 to 6795 participants (average 2036). All participants were older adults at baseline with a mean age ranging from 65 to 84.6 years (average 76 years). Gait speed was assessed using a stopwatch in most studies other than the three which reported on cohorts that used a computerized walkway.^4,5,8^ The measuring distance ranged from 2.4 to 6 meters and most cohorts measured usual walking speed, although two studies measured maximum walking speed.^10,46^ Slow gait was defined as one standard deviation or more below age- and sex-matched means in the population in all but two cohorts, one of which classed as slow walkers all those walking <0.8 meters per second (or <0.66 meters per second if female less than 1.45m in height)^16^ and the other which classed as slow walkers all those in the lowest 20^th^ percentile of the cohort population.^47^ Subjective cognitive complaint was measured using different methods in different cohorts, such as the memory item from the 15-item Geriatric Depression Scale,^8,10,11,15,17,44,46^ the eight-item informant interview (AD8),^17^ the Clinical Dementia Rating scale,^17^ the 15-item Consortium to Establish a Registry for Alzheimer’s Disease (CERAD) questionnaire.^4^ Of note, studies on the EPIDOS cohort included in this review and meta-analysis used an objective measure of cognitive complaint, namely any incorrect responses on the Short Portable Mental Status Questionnaire (SPMSQ). Accordingly, results should be treated with caution.^9,13,15^ In other studies, a positive response from participants to a question such as “Is your memory worse than 10 years ago?” was sufficient.^12^

##### 2.3.3 Results of ROB assessments

Figure 2 contains a summary ROB of the pooled studies (top) as well as a traffic-light plot (bottom) showing the ROB of the individual studies, assessed against the nine domains. In the pooled summary, the domains with highest ROB were due to missing data, exposure variable measurement, confounding and generalizability. In the traffic-light plot, there was a high ROB in at least one domain in the majority (7/11) of studies, and all studies contained at least one domain judged as being of some concern. Overall, six studies had low ROB, four studies had some concerns, and one study had a high ROB.

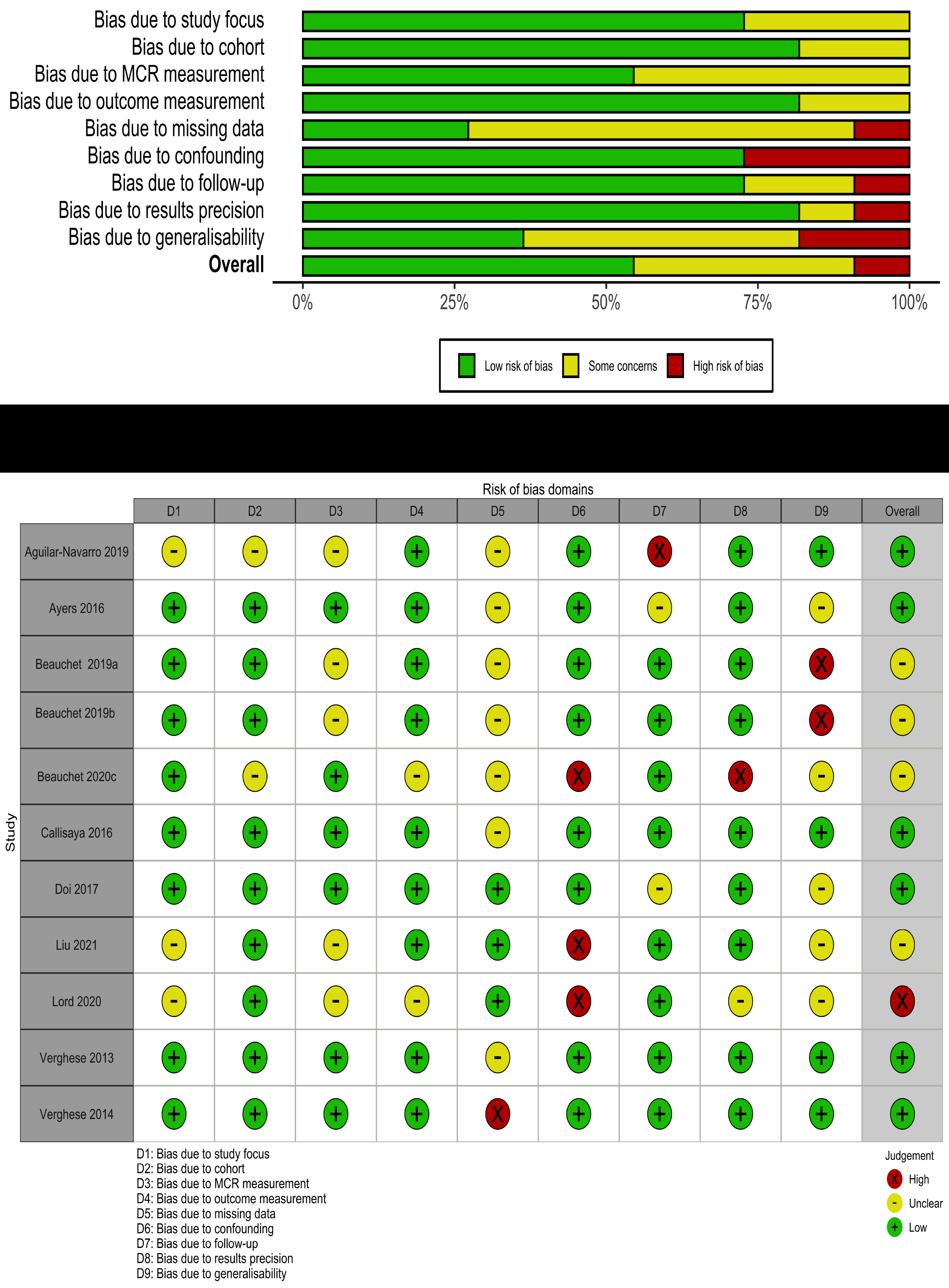

##### 2.3.4 Meta-analysis of health outcomes predicted by MCR

The meta-analysis findings are summarized in the forest plots and described in more detail below (figure 3).

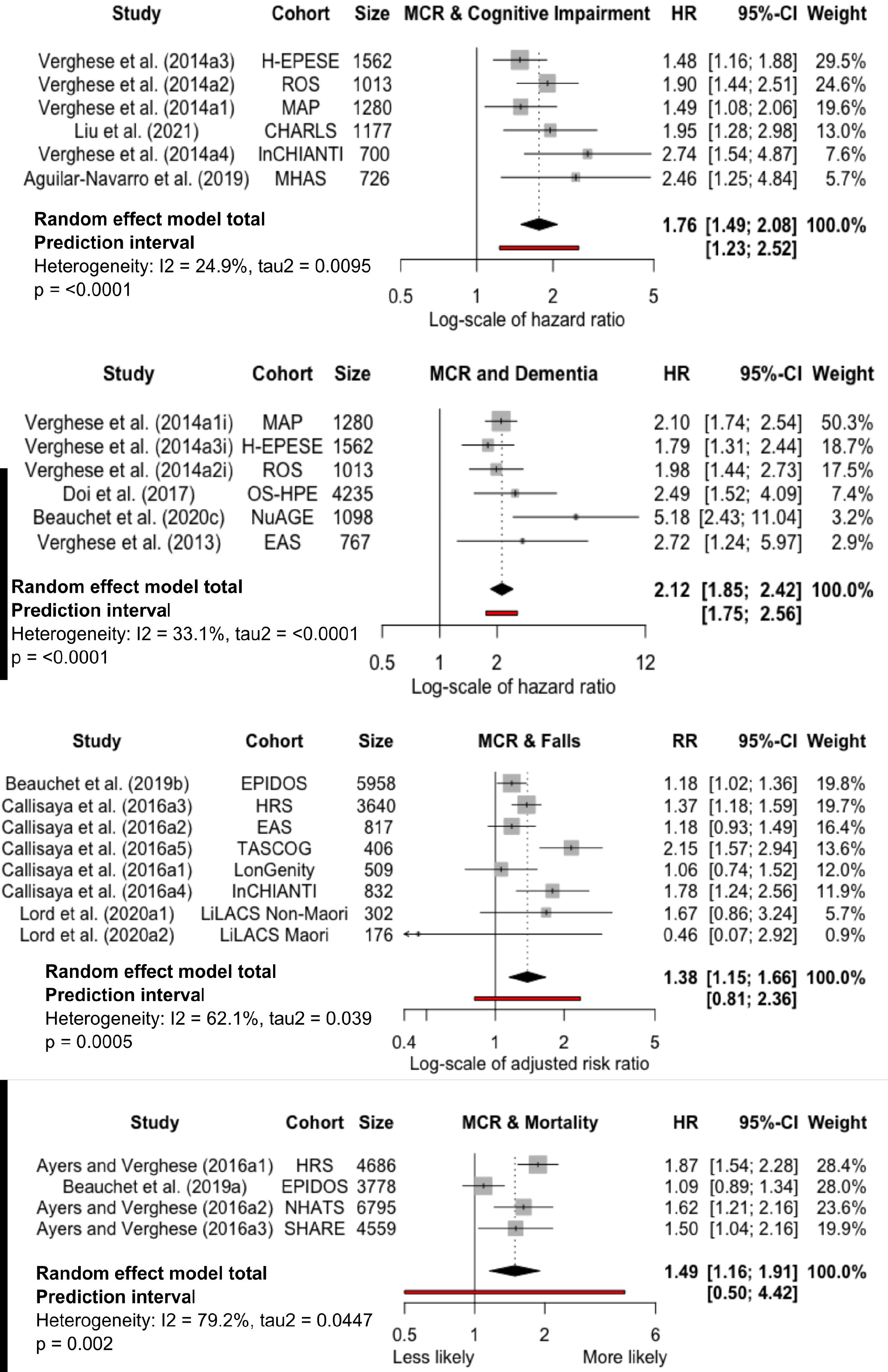

###### Dementia

We combined six cohorts from four studies that examined dementia as an outcome, totaling 9,955 participants.^4,5,11,45^ Four cohorts were from two studies with a low ROB^4,5^ and two were from studies with a moderate risk of overall bias.^11,45^ Individuals with MCR at baseline were over twice as likely to develop dementia compared to those without MCR, over an average follow-up of 4.3 years (aHR 2.12, 95%CI 1.85-2.42, p<0.0001, I^2^=33.1%, tau^2^=<0.0001, prediction interval 1.75-2.56).

###### Cognitive Impairment

Six cohorts from three studies, including a total of 6,458 participants, reported on cognitive impairment as an outcome. Five cohorts were from two studies with a low risk of overall bias^5,16^ and one^47^ with a moderate risk of overall bias. Those with MCR at baseline were at an increased risk of developing cognitive impairment compared to those without MCR, over an average follow-up of 5.6 years (aHR 1.76, 95%CI 1.49-2.08, p<0.0001, I^2^=24.9%, tau^2^=0.0095, prediction interval 1.23-2.52).

###### Falls

We combined eight cohorts from three studies, including a total of 12,640 participants with falls as an outcome. Two cohorts were from one study with a high risk of overall bias, mainly due to this study’s small sample size and method of dealing with confounders.^10^ These cohorts were weighted in the meta-analysis in accordance with their results imprecision, so their effect on the overall result was minimal (combined weighting of 6.3%). One cohort was from a study with a moderate ROB,^9^ while the other five cohorts were from a study with a low ROB.^8^ Those with MCR at baseline were at a 38% relative risk increase of falls compared to those without MCR (aRR 1.38, 95%CI 1.15-1.66, p=0.0005, I^2^=62.1%, tau^2^=0.039, prediction interval 0.91-2.36). The average follow-up of all cohorts was 1.4 years. A subgroup analysis excluding the two cohorts with a high risk of overall bias (total n=478 participants) made no difference to the effect measure of prognostic ability for MCR at baseline with falls on follow-up (aRR 1.38, 95%CI 1.14-1.69, p=0.0013, I^2^=70.2%, tau^2^=0.043, prediction interval 0.73-2.63).

###### Mortality

Four cohorts from two studies, including 19,818 participants, reported on mortality as an outcome. Three of these came from a study with a low risk of overall bias,^12^ with the other study^13^ having a moderate risk of overall bias. Those with MCR at baseline were at an increased risk of mortality compared to those without MCR, over an average follow-up of 5.3 years (aHR 1.49, 95%CI 1.16-1.91, p<0.0001, I^2^=79.2%, tau^2^=0.0477, prediction interval 0.50-4.42).

##### 2.3.5 Bias of studies or outcomes included in the meta-analysis

On visual inspection of the funnel plots, both the dementia and cognitive impairment outcomes appear to be missing smaller studies with a smaller effect size (figure 4). As there are fewer than ten studies for each outcome, any test for funnel plot asymmetry will have low power for distinguishing chance from real asymmetry.^40^ Nonetheless, the results from Egger’s test did not support asymmetry for dementia (p=0.15), cognitive impairment (p=0.06), falls (p=0.69) or mortality (p=0.54).

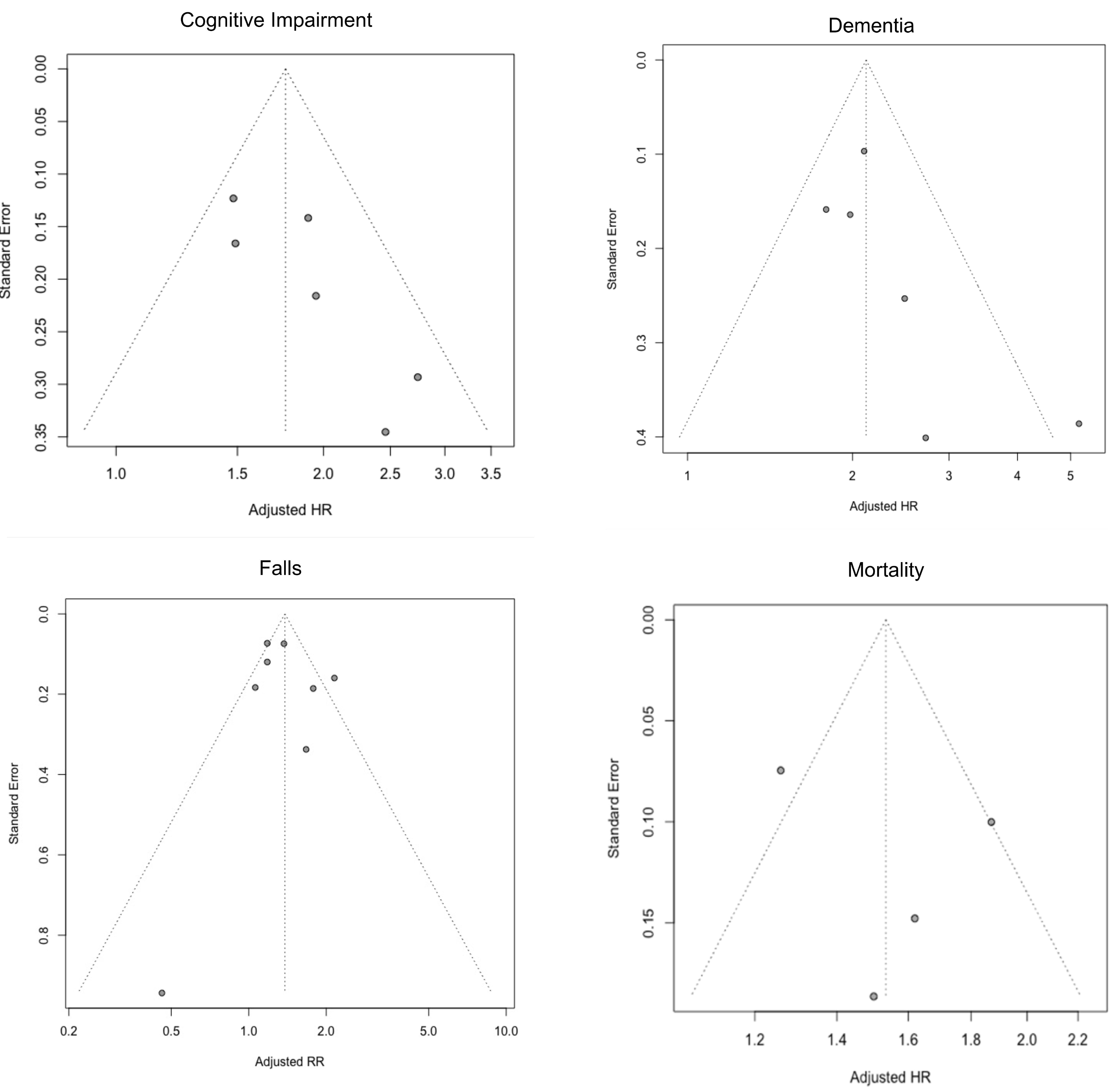

There was no evidence of selective non-reporting of results from any of the studies included in this review or meta-analysis.

### 3. Further detail on methods and results

#### 3.1 Further methodological details

##### 3.1.2 Quality assessment of included studies

Prior to ROB assessment, two authors (DM and AC) independently evaluated the methodological quality of the selected studies using a customized quality assessment tool that builds on the 14 criteria of the Cochrane recommended National Institute for Health (NIH) Quality Assessment Tool for Observational Cohort and Cross-Sectional Studies.^49^ An additional 15 criteria from the Strengthening The Reporting of Observational Studies in Epidemiology (STROBE) guidelines^50^ were added to our tool, resulting in a total of 29 criteria by which each study was assessed (supplementary table 4). Two authors independently rated each study quality overall as “high” if most criteria were met and there was little risk of bias (ROB), “satisfactory” if most criteria were met with some flaws in the study, or “low” when most criteria were not met, and/or there were significant flaws relating to key aspects of study design. Any discrepancies in judgements were resolved by discussion to reach a consensus. There were no discrepancies in our independent overall impression of each study. Our approach fits with that recommended by the Cochrane Handbook v6.2.^40^

In addition to this detailed quality assessment, we performed a more succinct ROB assessment as described in section 1.5.5. From the criteria in our quality assessment tool, we selected those most relevant to ROB: (i) bias due to lack of study focus; (ii) bias arising from cohort used; (iii) bias due to MCR measurement; (iv) bias due to outcome measurement; (v) bias due to missing data; (vi) bias due to confounding; (vii) bias due to follow-up; (viii) bias due to results precision; (ix) bias due to lack of generalizability. These criteria are more appropriate when assessing ROB of studies describing a syndrome such as MCR than the recently developed Prediction model Risk Of Bias ASsessment Tool (PROBAST) tool,^51^ which assesses multivariable prediction models. The risk of bias plots were created in the coding language R using the “robvis (0.3.0.9)” package.^52^ The code syntax is openly available on GitHub (https://github.com/d-mullin?tab=repositories).

##### 3.1.2 Assessment of certainty in the body of evidence

We assessed the certainty of the body of evidence for MCR as a predictor of each outcome in the meta-analysis against the GRADE considerations adapted for prognosis research as recommended by Huguet^43^, see section 1.6.5. We graded the certainty in the results for MCR as a predictor of dementia, cognitive impairment, falls, and mortality as low (see table 2). This low certainty is typical for prognosis research using even sound observational studies.^40^ The seven GRADE criteria considered, and our impressions for each, are described in supplementary box 2 of the appendix.

**Table 2:**
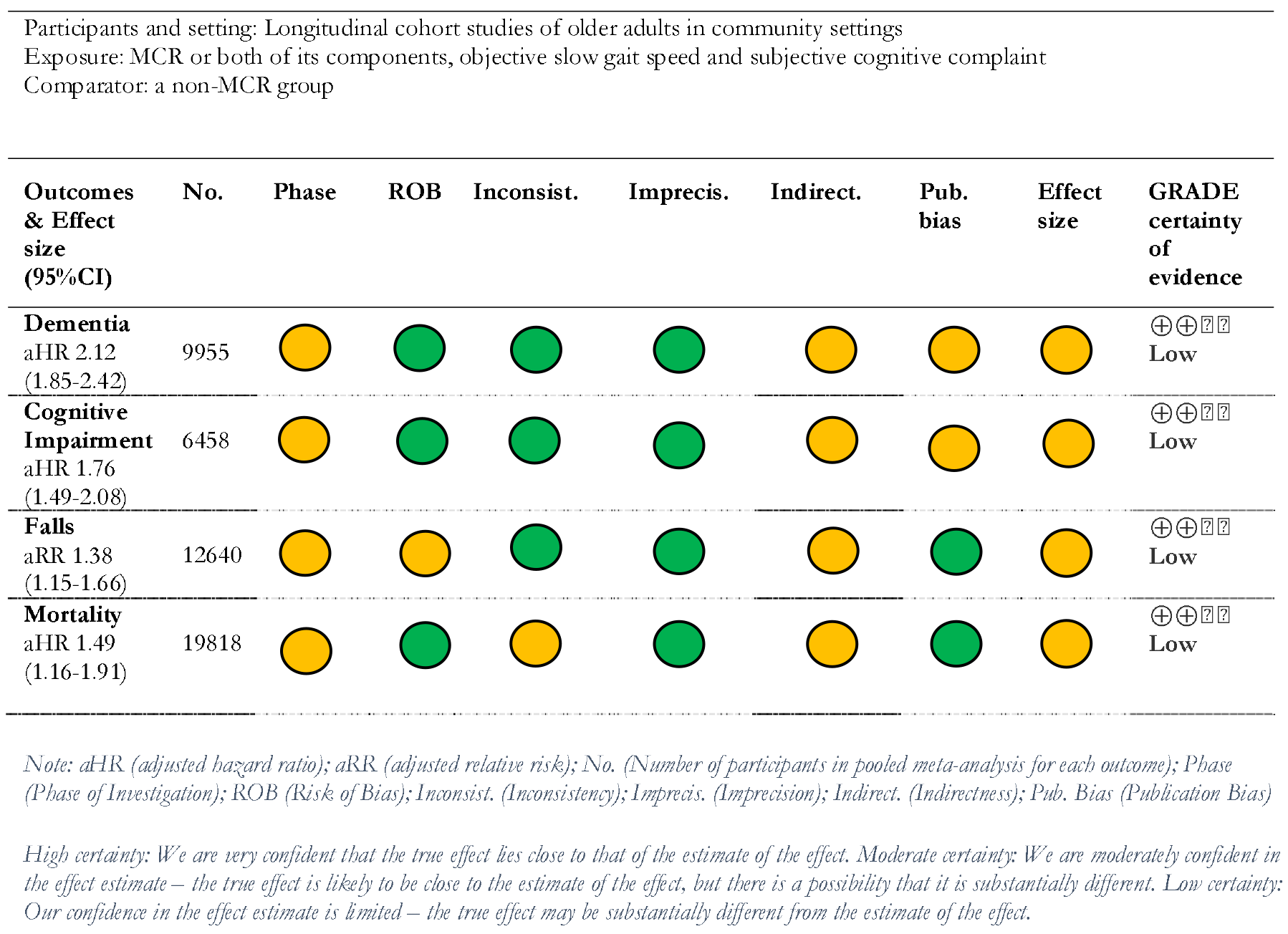
GRADE assessment of certainty in the body of evidence for MCR as a predictor of each health outcome

### 3.2 Further results

#### 3.2.1 Studies excluded from meta-analysis

A further two eligible studies^15,46^ reported on MCR as a predictor of dementia and two more as a predictor of cognitive impairment^17,44^ but, due to how the effect size was reported in each, (e.g., reporting OR rather than HR) we were unable to include them in our meta-analysis. Despite correspondence with study authors, we did not obtain the data necessary to allow calculation of the effect measure used in our meta-analysis. Reassuringly, these excluded studies share similar characteristics and direction of effect size to those included in the meta-analysis (table 3). They were judged as having either a moderate or high ROB. The effect precision was poor in three of the studies and not reported in the other, so if it had been possible to include these in the meta-analysis, they would have had a relatively small weighting and therefore, a minimal impact on the overall result.

**Table 3.**
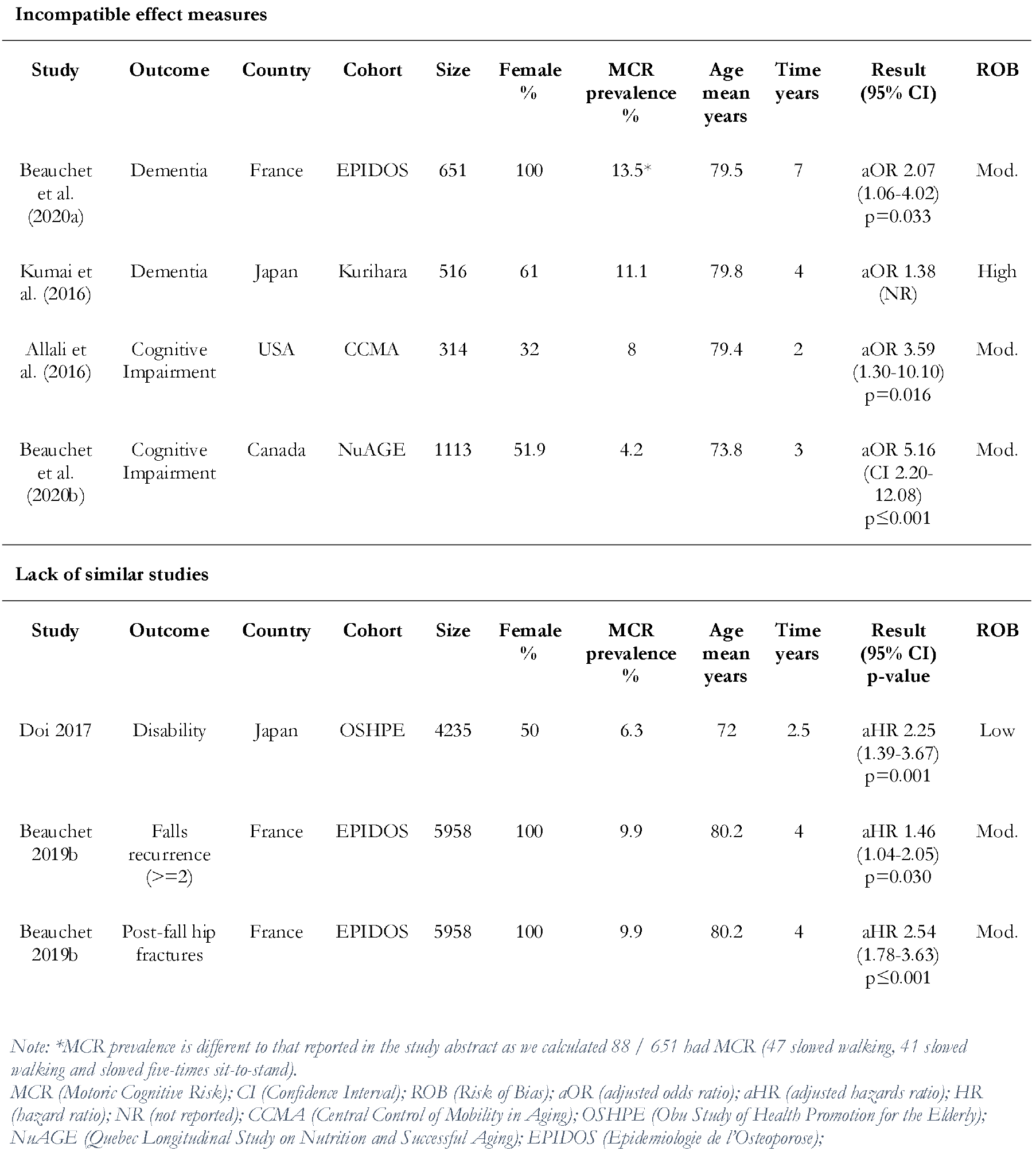
Characteristics of studies excluded from meta-analysis due to incompatible effect measures or lack of similar studies

MCR at baseline was reported to predict other health outcomes on follow-up, but there were not enough different cohorts reporting on the same outcomes to allow for meta-analysis. One study found that MCR was a predictor of incident disability after 2.5 years (aHR 2.25 (1.39-3.67) p=0.001).^11^ Disability was defined as a primary care doctor evaluated need for new long-term care insurance certification. Another study, already included in our meta-analysis for falls at one year, also reported on MCR as a predictor of recurrent falls, defined as two or more falls, (aHR 1.46 (1.04-2.05) p=0.030) and of post-falls hip fractures (aHR 2.54 (1.78-3.63) p≤0.001).^9^

## Research in Context

### Systematic review

We systematically retrieved and reviewed the literature using traditional sources and correspondence with authors. While the predictive value of MCR for cognitive impairment, dementia, falls, and mortality have been reported in sufficient cohorts to allow for meta-analysis, other outcomes such as disability were not, so were reported in narrative form.

### Interpretation

Our findings led to an integrated hypothesis describing the pathophysiology of MCR. This hypothesis is consistent with epidemiological, imaging, and genetic findings currently in the public domain.

### Future directions

This manuscript proposes a framework for the generation of new hypotheses and describes a roadmap to validate these hypotheses. Examples include: (a) clarifying the definition of subjective cognitive complaint; (b) longitudinal studies with biomarkers and pathological analysis; (c) genome wide association studies of MCR to identify genetic polymorphisms and potential treatment targets of interest.

## Supporting information

Highlights

## Data Availability

All data produced in the present study are available upon reasonable request to the authors

## Acknowledgements

Ruth Jenkins, academic support librarian at the University of Edinburgh, helped with search terms and recommending databases. Dr Sue Lord, Lecturer at the Auckland University of Technology, New Zealand, supplied raw data from her study to allow for calculation of appropriate effect measure.

---

Table 1 – Characteristics of studies

Table 2 – GRADE assessment of certainty in results

Table 3 – Excluded studies

