## Supplementary material for "PREPRINT: Mechanisms of Motoric Cognitive Risk – hypotheses based on a systematic review and meta-analysis of longitudinal cohort studies of older adults": Highlights

- Motoric Cognitive Risk (MCR) is a syndrome combining slow gait and self-reported cognitive complaints
- MCR is prognostic of incident dementia and other major causes of morbidity in older age
- Meta-analysis found people with MCR were at an increased risk of cognitive impairment and dementia
- They were also at increased risk of falls and mortality
- Based on the literature, possible mechanisms underlying MCR were discussed
