## Supplementary Material for "PREPRINT: Mechanisms of Motoric Cognitive Risk – hypotheses based on a systematic review and meta-analysis of longitudinal cohort studies of older adults"

Alternate Abstract – in keeping with those published in *Alz* *& Dem* for theoretical papers

Supplementary Table 1 – Search strategy

Supplementary Table 2 – Data extraction tool

Supplementary Table 3 – Summary of included studies

Supplementary Table 4 – Quality assessment tool

Supplementary Box 1 – PICOT system for review question

Supplementary Box 2 – GRADE Criteria for assessment of certainty in results

**Alternate Abstract**

**Objective**

We aimed to refine the hypothesis that Motoric Cognitive Risk (MCR), a syndrome combining measured slow gait speed and self-reported cognitive complaints, is prognostic of incident dementia and other major causes of morbidity in older age. We propose mechanisms on the relationship between motor and cognitive function and describe a roadmap to validate these hypotheses.

**Background**

MCR is being used to identify older adults at high risk of developing dementia. Yet, uncertainty exists regarding the prognostic value of MCR for dementia and other adverse health outcomes in ageing, such as cognitive impairment, falls, and mortality.

**Updated Hypothesis**

We hypothesize that participants with MCR are at an increased risk of developing cognitive impairment, dementia, falls, and mortality. We systematically searched major electronic databases from inception to August 2021 for original longitudinal cohort studies of adults aged ≥60 years that compared an MCR group to a non-MCR group with any health outcome. Fifteen cohorts were combined by meta-analysis using a restricted maximum-likelihood estimator reporting on four health outcomes: cognitive impairment (n=6,458), dementia (n=9,955), falls (n=12,640), and mortality (n=19,818). Risk of bias was deemed low-to-moderate in 13/15 cohorts. Our meta-analysis found that participants with MCR were at an increased risk of cognitive impairment (adjusted hazard ratio [aHR] 1.76, 95%CI 1.49-2.08; I^2^=24.9%), dementia (aHR 2.12, 1.85-2.42; 33.1%), falls (adjusted Relative Risk 1.38, 1.15-1.66; 62.1%), and mortality (aHR 1.49, 1.16-1.91; 79.2%).

**Major Challenges for the Hypothesis**

We hypothesize that cognitive impairment, dementia, and falls could share underlying pathologies with MCR, ultimately resulting in increased mortality. MCR is quick, cheap, and practical to measure in clinic and could prove to be a valuable screening tool to inform referral to specialist services. The range of methods for diagnosing subjective cognitive complaint highlights the need for a consistent definition of MCR in future studies. Longitudinal studies with biomarkers and pathological validation will help determine the discriminatory ability of MCR to predict different subtypes of dementia.

**Linkage to Other Major Theories**

These meta-analyses findings support the theory of interacting motor-cognitive networks and raise the exciting possibility that MCR may be an early noninvasive biomarker for the major causes of morbidity and mortality of old age, including dementia.

Supplementary Table 1 – Search strategy

Database: Ovid MEDLINE(R) <1946 to August Week 1 2021>

Search Strategy:

--------------------------------------------------------------------------------

1	gait/ or gait analysis/ or walking speed/ (29635)

2	Gait Disorders, Neurologic/ (6748)

3	Walking/ (33564)

4	(speed or pace or velocity or time*).mp. [mp=title, abstract, original title, name of substance word, subject heading word, floating sub-heading word, keyword heading word, organism supplementary concept word, protocol supplementary concept word, rare disease supplementary concept word, unique identifier, synonyms] (4293037)

5	1 or 2 or 3 (59712)

6	4 and 5 (28287)

7	Cognition Disorders/ or Cognition/ (156587)

8	Memory, Long-Term/ or Memory/ or Memory Disorders/ or Memory, Short-Term/ or Memory, Episodic/ (110781)

9	7 or 8 (245574)

10	(subjective or self-report* or self-declar* or self-identif* or patient-report* or self-perceive*).mp. [mp=title, abstract, original title, name of substance word, subject heading word, floating sub-heading word, keyword heading word, organism supplementary concept word, protocol supplementary concept word, rare disease supplementary concept word, unique identifier, synonyms] (254232)

11	6 and 9 (1132)

12	10 and 11 (101)

13	"motoric cognitive risk".mp. [mp=title, abstract, original title, name of substance word, subject heading word, floating sub-heading word, keyword heading word, organism supplementary concept word, protocol supplementary concept word, rare disease supplementary concept word, unique identifier, synonyms] (40)

14	12 or 13 (133)

*Note: This search strategy was adjusted as appropriate for each database searched*

Supplementary Table 2 – Data extraction tool

| **Extracted information** | **Included details** |
| --- | --- |
| General information | Author, title, journal, year, related or duplicate publications |
| Source of data | Cohort, country |
| Sample size | Number of participants and number of outcomes/events |
| Participant information | Age, sex, proportion with MCR |
| MCR | Definition and method of measurement of MCR: slow gait measurement protocol, average gait speed, subjective cognitive complaint measurement method |
| Outcomes to be predicted | Definition and method of measurement of outcome; time of outcome ascertainment, or summary of duration of follow‐up |
| Adjustment for other prognostic factors (covariates) | List of all the covariates that were adjusted for in any regression model |
| Reported results | We recorded the adjusted and, where available, unadjusted model results of any health outcome result incidence, whether reported as hazard ratio (HR), odds ratio (OR) or relative risk (RR), and their corresponding 95% confidence intervals (CI) and p-values, if available. |
| Funding Source | We recorded the funding source of each study |

**Supplementary Table 3** Summary of included studies of the association between motoric cognitive risk and negative health outcomes

| **MCR as a predictor of Dementia** | | | | | | | | | | | | | | |
| --- | --- | --- | --- | --- | --- | --- | --- | --- | --- | --- | --- | --- | --- | --- |
| **Study** | **Country - Cohort** | **Size – MCR cases (MCR prevalence)** | **Female %** | **Age mean years** | | **Measurement of MCR** | | **Measurement of outcome** | **Duration years** | **Covariates included in most-adjusted model** | | | **Result (95% CI)** | **Risk of Bias** |
|  |  |  |  |  | | **Gait** | **SCC** |  |  |  | | |  |  |
| Doi et al. (2017) | Japan - OSHPE | 4235 – 265 (6.3%) | 50 | 72 | | 2.4m usual pace stopwatch. 2m initial acceleration and terminal deceleration space | "Do you feel you have more problems with memory than most?" | Incident cases of dementia were identified from monthly insurance data based on ICD-10 diagnosis from doctor. | 2.5 | Less education, diabetes, obesity, sedentariness, depression, drinking alcohol, CVD, fall, higher medication numbers | | | 2.49 (1.52, 4.1) | Low |
| Verghese et al. (2013) | USA - EAS | 767 – 52 (6.8%) | 60 | 79.9 | | Usual pace - GAITRite, two practice trials over 4.60m then assessed over 6.10m. | 15-item CERAD questionnaire. Also, study clinicians' observations during clinical interview | Case conference by clinicians who diagnosed dementia based on DSM - IV criteria | 3.1 | Age, sex, and education, Blessed test score, medical illness index (depression, diabetes, heart failure, hypertension, angina, MI, strokes, Parkinson’s disease, chronic lung disease, arthritis) | | | 2.72 (1.24, 5.97) | Low |
| Verghese et al. (2014a1i) | USA - MAP | 1280 – 166 (13%) | 57 | 79.9 | | 8 ft timed | Self-report questionnaire | DSM IIIR | 5.1 | Age, sex, education, cohort source, baseline MMSE scores, and vascular disease | | | 2.1 (1.43, 2.09) | Low |
| Verghese et al. (2014a2i) | USA - ROS | 1013 – 132 (13%) | 57 | 75.1 | | 8 ft timed | Self-report questionnaire | DSM IIIR | 9.3 | Age, sex, education, cohort source, baseline MMSE scores, and vascular disease | | | 1.98 (1.44, 2.74) | Low |
| Verghese et al. (2014a3i) | USA - H-EPESE | 1562 – 141 (9%) | 57 | 72.3 | | 9 ft timed | IADL questionnaire | Clinical diagnosis | 6 | Age, sex, education, cohort source, baseline MMSE scores, and vascular disease | | | 1.79 (1.31, 2.44) | Low |
| Beauchet et al. (2020c) | Canada - NuAGE | 1098 – 46 (4.2%) | 50.7 | 73.8 | | 4-metre distance at their usual pace. Best of two. 1m initial acceleration space | GDS - more problems with memory than most? | Dementia - Modified Mini-Mental State (≤79/100) test and Instrumental Activity Daily Living scale (≤6/8) score values. | 3 | Age, sex, baseline 3MS score, abnormal IADL score, depression | | | 5.18 (2.43, 11.03) | Mod. |
| **MCR as a predictor of Cognitive Impairment** | | | | | | | | | | | | | | |
| **Study** | **Country - Cohort** | **Size – MCR cases (MCR prevalence)** | **Female %** | **Age mean years** | | **Measurement of MCR** | | **Measurement of outcome** | | **Duration years** | **Covariates included in most-adjusted model** | | **Result (95% CI)** | **Risk of Bias** |
|  |  |  |  |  | | **Gait** | **SCC** |  | |  |  | |  |  |
| Aguilar-Navarro et al. (2019) | Mexico - MHAS | 726 – 104 (14.3%) | 54 | 69.8 | | Walk usual pace 4 m twice then averaged | Responding “inadequate” or “poor” to the question: Compared to the last two years, would you say your memory is? | Total score in the cognitive test (CCCE) equal or less than -1.5 standard deviations and an IQCODE greater than 3.4 points | | 2.9 | Age, education, history of diabetes, hypertension, falls and depression | | 2.46 (1.25, 4.84) p=0.009 | Low |
| Verghese et al. (2014a1) | USA - MAP | 1280 – 166 (13%) | 57 | 79.9 | | 8 ft timed | Self-report questionnaire | DSM IIIR | | 5.1 | Age, sex, education, cohort source, baseline MMSE scores, and vascular disease | | 1.49 (1.08, 2.07) p=0.015 | Low |
| Verghese et al. (2014a2) | USA - ROS | 1013 – 132 (13%) | 57 | 75.1 | | 8 ft timed | Self-report questionnaire | DSM IIIR | | 9.3 | Age, sex, education, cohort source, baseline MMSE scores, and vascular disease | | 1.9 (1.44, 2.51) p=0.001 | Low |
| Verghese et al. (2014a3) | USA - H-EPESE | 1562 – 141 (9%) | 57 | 72.3 | | 9 ft timed | IADL questionnaire | Clinical diagnosis | | 6 | Age, sex, education, cohort source, baseline MMSE scores, and vascular disease | | 1.48 (1.16, 1.88) p=0.002 | Low |
| Verghese et al. (2014a4) | Italy - InCHIANTI | 700 – 56 (8%) | 57 | 74.1 | | 4 m timed | WHO disability scale | DSM IV | | 7.2 | Age, sex, education, cohort source, baseline MMSE scores, and vascular disease | | 2.74 (1.54, 4.86) p=0.001 | Low |
| Liu et al. (2021) | China - CHARLS | 1177 – 79 (6.7%) | 45.3 | 65 | | 2.5 m timed | Answered “poor” to the following survey item: “How would you rate your memory at the present time? Would you say it is excellent, very good, good, fair, or poor? | The lowest 10% of the distribution of global cognition during follow-ups. Global cognition was scored using the summation of the episodic memory and mental intactness scores, which ranges from 0 to 31. | | 4 | Age, sex, education, baseline cognition | | 1.95 (1.21, 2.82) | Mod. |
| **MCR as a predictor of Falls** | | | | | | | | | | | | | | |
| **Study** | **Country - Cohort** | **Size – MCR cases (MCR prevalence)** | **Female %** | | **Age mean years** | **Measurement of MCR** | | **Measurement of outcome** | | **Duration years** | **Covariates included in most-adjusted model** | | **Result (95% CI)** | **Risk of Bias** |
|  |  |  |  | |  | **Gait** | **SCC** |  | |  |  | |  |  |
| Callisaya et al. (2016a1) | USA - LonGenity | 509 – 56 (11%) | 52.3 | | 75 | GAITRite | GDS/self-report | "any falls in last 12 months" | | 1.5 | Age and sex | | 1.06 (0.74, 1.52) | Low |
| Callisaya et al. (2016a2) | USA - EAS | 817 – 99 (12.1%) | 61.6 | | 79.7 | GAITRite | GDS/self-report | "any falls in the past 2 months / past year" | | 1 | Age and sex | | 1.18 (0.93, 1.49) | Low |
| Callisaya et al. (2016a3) | USA - HRS | 3640 – 244 (6.7%) | 56.9 | | 74.4 | 2.5m walk normal pace | Self-report | "fallen down in the last two years" | | 2 | Age and sex | | 1.37 (1.18, 1.58) | Low |
| Callisaya et al. (2016a4) | Italy - InCHIANTI | 832 – 57 (6.9%) | 55.2 | | 73.5 | 4m walk | Disability scale | "Did you ever fall down in the past months" | | 3 | Age and sex | | 1.78 (1.23, 2.55) | Low |
| Callisaya et al. (2016a5) | Australia - TASCOG | 406 – 7 (1.7%) | 43.1 | | 72 | GAITRite | GDS/self-report | "Have you had any falls in months of _" (2 monthly questionnaire over 1 year) | | 1 | Age and sex | | 2.15 (1.57, 2.94) | Low |
| Beauchet et al. (2019b) | France - EPIDOS | 5958 - 590 (9.9%) | 100 | | 80.2 | 6 m at usual pace. No acceleration phase but a 2m terminal deceleration phase. First trial of three | SCC was considered when there were one or two incorrect SPMSQ answers- | Self-report retrospectively at annual assessment | | 1 | Not adjusted | | 1.18 (1.02, 1.36) | Mod. |
| Lord et al. (2020a1) | New Zealand - LiLACS NZ Non-Maori | 302 - 6 (1.9%) | 54.1 | | 84.6 | Fastest speed of two trials over a 3 m distance | GDS/self-report | Self-report retrospectively at annual assessment | | 1 | Age and sex | | 1.67 (0.86, 3.23) | High |
| Lord et al. (2020a2) | New Zealand - LiLACS NZ Maori | 176 - 8 (4.3%) | 58.2 | | 82.6 | Fastest speed of two trials over a 3 m distance | GDS/self-report | Self-report retrospectively at annual assessment | | 1 | Age and sex | | 0.46 (0.07, 2.83) | High |
| **MCR as a predictor of Mortality** | | | | | | | | | | | | | | |
| **Study** | **Country - Cohort** | **Size – MCR cases (MCR prevalence)** | **Female %** | **Age mean years** | | **Measurement of MCR** | | **Measurement of outcome** | **Duration years** | **Covariates included in most-adjusted model** | | **Result (95% CI)** | | **Risk of Bias** |
|  |  |  |  |  | | **Gait** | **SCC** |  |  |  | |  | |  |
| Ayers and Verghese (2016a1) | USA - HRS | 4686 - 375 (8%) | 56.4 | 74.7 | | 2.5m walk normal pace | Self-report questionnaire | Family/friend report. Reported death dates were confirmed through the social security death index and insight databases, and linked to the national death index | 6 | Age, sex, and education | | 1.87 (1.54, 2.28) | | Low |
| Ayers and Verghese (2016a2) | USA - NHATS | 6795 - 435 (6.4%) | 57.2 | 77.3 | | 3m walk normal pace | Self-report questionnaire | Proxy-respondent report. | 2.1 | Age, sex, and education | | 1.62 (1.21, 2.16) | | Low |
| Ayers and Verghese (2016a3) | 11 European countries - SHARE | 4559 - 319 (7%) | 55.7 | 81.7 | | 2.5m walk normal pace | Self-report IADL questionnaire | Proxy-respondent report. | 4 | Age, sex, and education | | 1.5 (1.04, 2.16) | | Low |
| Beauchet et al. (2019a) | France - EPIDOS | 3778 - 382 (10.1%) | 100 | 80.5 | | 6 m at usual pace. No acceleration phase but a 2m terminal deceleration phase. First trial of three | SCC was considered when there were one or two incorrect SPMSQ answers- | Deaths were prospectively recorded using mail, phone calls, questionnaires and/or the French national death registry | 5 | Age, education, place of living, BMI, number of drugs taken daily, use of psychoactive drugs, history of depression, physical activity level, history of falls in the past year, cardiovascular risk factors (obesity, smoking, hypertension, angina, diabetes, and history of CVA)) and the recruitment centre | | 1.09 (0.89, 1.34) p=0.401 | | Mod. |

Note: Results are adjusted hazard ratios except for those cohorts with falls as the outcome, which are adjusted relative risk. ROB gradings – 1 (low), 2 (some concern), 3 (high). MCR (Motoric Cognitive Risk); CI (Confidence Interval); Risk of Bias (ROB); OSHPE (Obu Study of Health Promotion for the Elderly); EAS (Einstein Aging Study); MAP (Memory and Aging project); ROS (Religious Order Study); H-EPESE (Hispanic Established Population for Epidemiological studies of the Elderly); NuAGE (Quebec Longitudinal Study on Nutrition and Successful Aging); MHAS (Mexican Health and Aging Study); InCHIANTI (Invecchiare in Chianti)I; CHARLS (China Health and Retirement Survey); LonGenity; TASCOG (The Tasmanian Study of Cognition and Gait); EPIDOS (Epidemiologie de l’Osteoporose); LiLACS NZ (Life and Living in Advanced Age, a Cohort Study in New Zealand); NHATS (National Health and Aging Trends Study); SHARE (Survey of Health, Ageing and Retirement in Europe)

Slow gait was defined as ≥1 standard deviation above age- and sex-matched mean in all studies except Aguilar-Navarro^16^ (slower than 0.8 metres per second in both men and women; except for women with a height <1.45 m in which the cut-off value was >0.66 metres per second) and Liu^47^ (lowest 20^th^ percentile of population distribution adjusted for sex and height)

Supplementary Table 4 – Quality assessment tool.

Adapted from National Institute for Health (NIH) Quality Assessment Tool for Observational Cohort and Cross-Sectional Studies^49^ and Strengthening the Reporting of Observational Studies in Epidemiology (STROBE) guidelines.^50^ Note: Each criterion was answered as “yes,” “no,” “cannot determine,” “not applicable” or “not reported”. Two authors independently rated each study overall as “high” if most criteria were met, “satisfactory” if most criteria were met with some flaws in the study, or “low” when most criteria were not met, and/or there were significant flaws relating to key aspects of study design.

**Abstract**

1. Is the abstract an informative and balanced summary of what was done and what was found?

**Introduction**

2. Objectives - do authors state specific objectives, including any prespecified hypotheses?

**Methods**

3. Setting - Do authors describe the setting, locations, and relevant dates, including periods of recruitment, exposure, follow-up, and data collection - or refer to where these are clearly described? Was the timeframe sufficient so that one could reasonably expect to see an association between exposure and outcome, if it existed?
4. Participants - Do authors give the eligibility criteria, and the sources and methods of selection of participants. Do they describe methods of follow-up (if longitudinal)?
5. Participants - Do authors include a flowchart of sample selection?
6. Were the exposure/independent variables clearly defined, valid, reliable, and implemented consistently across all study participants, especially if there is more than one group? Do they give diagnostic criteria, if applicable?
7. Were the outcome/dependent variables clearly defined, valid, reliable, and implemented consistently across all study participants, especially if there is more than one group? Do they give diagnostic criteria, if applicable?
8. Were potential confounding variables clearly identified, or included in limitations section if not? Were effect modifiers and mediators described?
9. Bias - do authors describe any efforts to address potential sources of bias?
10. Study size - do authors explain how the study size was arrived at?
11. Quantitative variables - do authors explain how quantitative variables were handled in the analyses? If applicable, do they describe which groupings were chosen and why?

**Statistical methods - do the authors?:**

12. Describe all statistical methods, including those used to control for confounding
13. Explain how missing data were addressed
14. Cohort study—If applicable, explain how loss to follow-up was addressed
15. Describe any sensitivity analyses

**Results**

16. Participants - (a) Do authors report numbers of individuals at each stage of study—eg numbers potentially eligible, examined for eligibility, confirmed eligible, included in the study, completing follow-up, and analysed.
17. Participants - (b) Do authors give reasons for non-participation at each stage? Was the participation rate of eligible persons at least 50%?
18. Was loss to follow-up after baseline 20% or less?
19. Outcome data - do authors report numbers of outcome events or summary measures (over time, if longitudinal)?
20. Do the authors give unadjusted estimates and, if applicable, confounder-adjusted estimates and their precision (e.g., 95% confidence interval)? Make clear which confounders were adjusted for and why they were included?
21. Do the authors report category boundaries when continuous variables were categorized?
22. Do the authors, if applicable, consider translating estimates of relative risk into absolute risk for a meaningful time period?
23. Other analyses - do authors report other analyses done—e.g. analyses of subgroups and interactions, and sensitivity analyses?
24. Do you believe the results?

**Discussion**

25. Key results - Do authors summarise key results with reference to study objectives
26. Limitations - Do authors discuss limitations of the study, considering sources of potential bias or imprecision. Consider also if they discuss both direction and magnitude of any potential bias?
27. Interpretation - Do authors give a cautious overall interpretation of results considering objectives, limitations, multiplicity of analyses, results from similar studies, and other relevant evidence?
28. Generalisability - Do authors discuss the generalisability (external validity) of the study results?

**Funding**

29. Is there a statement of funding or conflicts of interest?

**Overall rating** (see explanations below)

High: Majority of criteria met. Little or no risk of bias

Satisfactory: Most criteria met. Some flaws in the study with an associated risk of bias

Low: Either most criteria not met, or significant flaws relating to key aspects of study design

Number of yes answers - max is 29 (this is purely for checking overall rating scores match up, not for calculating overall rating)

Supplementary Box 1 PICOT criteria for review question

Population: Community-based adults aged ≥60 years.

Intervention: MCR or the MCR criteria of objective slow gait speed and subjective cognitive complaint, in the absence of significant functional impairment and dementia. Importantly, the term MCR did not have to be used. This allowed for consideration of all studies examining the phenotype of objective slow gait and subjective cognitive complaint, rather than just those using the term MCR, which was only coined in 2013.

Comparator: Included studies contained a comparator group who did not have MCR, or each of its component parts. We compared the most adjusted model of MCR to other (that is, comparator) prognostic factors such as age, sex, education, comorbidities.

Outcome: We included studies reporting any health outcome, including, but not limited to those used in the meta-analysis (i.e., cognitive impairment, dementia, falls and mortality). Studies were omitted from the meta-analysis if their method of reporting results did not allow for synthesis with other studies. All attempts were made to both obtain data directly from authors and to transform effect measures when data was not obtained from authors, but still some studies were omitted and are listed in the meta-analysis section.

Type of study: We included longitudinal, observational cohort studies.

Timing and setting: Recruitment from primary, secondary, or community settings. Minimum of one year follow-up to allow time for outcomes to develop.

Authors of posters or abstracts were contacted directly to enquire if a full-text version had been published when one was not obvious in the literature search.

*Supplementary Box 2 GRADE assessment of certainty in our results*

1. Phase of investigation: We considered as high-quality evidence any phase 3 explanatory studies derived from custom-made cohort study designs that aimed to understand the pathway between MCR and the health outcome. We considered as moderate-quality evidence any phase 2 explanatory studies that aimed to confirm independent associations between MCR and the health outcome. We considered as low-quality evidence any phase 1 hypothesis generating explanatory studies that aim to identify associations between MCR and the health outcome. All the cohorts came from either phase 1 or phase 2 explanatory studies, leading to a downgrading of the level of evidence for all health outcomes.
2. ROB: This judgement was based on our expanded QUIPS tool, as described previously.
3. Inconsistency: We downgraded the evidence if associations between MCR and the health outcome were markedly heterogeneous (i.e., the reported aHR/aRR fell either side of 1 on a forest plot or if the I^2^ statistic was substantial (i.e., 75% or more)). We downgraded the evidence for MCR as a predictor of mortality as the I^2^ statistic was substantial at 79%.
4. Imprecision: We downgraded the evidence if there were insufficient numbers in the meta-analysis or if the confidence intervals were wide. There were enough cohorts with sufficiently narrow confidence intervals to generate adequately precise results on meta-analysis.
5. Indirectness: We downgraded the evidence if the participant population, the MCR criteria used and/or the outcomes investigated did not fully match with our review question. We downgraded the evidence if the participant population, the MCR criteria used and/or the outcomes investigated did not fully match with our review question.
6. Publication bias: We took the prudent default position of assuming that this prognosis research is affected by publication bias.^43^ Unless there was significant evidence to the contrary, such as a symmetrically distributed funnel plot of a large number of cohorts, we downgraded the evidence.
7. Effect size: We upgraded our confidence in the effect estimate if the effect size was moderate to large (e.g., aHR >2.5). We did not upgrade our confidence in the effect estimate for any health outcome.
